# The Wood equation allows consistent fitting of individual antibody responses profiles in Zika virus or SARS-CoV-2 infected patients

**DOI:** 10.1101/2020.12.16.20248157

**Authors:** J. Denis, A. Garnier, D. Claverie, F. De Laval, S. Attoumani, B. Tenebray, G.A. Durand, B. Coutard, I. Leparc-Goffart, JN. Tournier, S. Briolant, C. Badaut

## Abstract

Antibody kinetic curves obtained during a viral infection are often fitted using aggregated data from patients, hiding the heterogeneity of patient responses. The Wood equation makes it possible to establish the link between an individual’s kinetic profile and the disease, which may be helpful in identifying and studying clusters.

## Introduction

Viral infections are followed by an immune response, generally leading to increased antibody levels. The severity of the disease following infection with severe acute respiratory syndrome coronavirus 2 (SARS-CoV-2) can be related to the antibody kinetics [1]. Several studies have also shown a difference in the kinetic profiles of anti-Zika virus (ZIKV) antibodies, depending on whether or not there was a pre-dengue infection [2].

IgG/IgM kinetic profiles may allow investigation of the link between a humoral immune response and its involvement in the severity of the resulting disease until its disappearance or clearance of the infecting virus. The immune response is characterized by the amplitude of the antibody response and the day on which they become detectable, their concentration is maximal, and they become undetectable. These data are difficult to obtain. Furthermore, data from various patients are often incomplete and aggregated to obtain an average kinetic curve that does not account for the heterogeneity of the humoral immune response of each patient. Here, we propose using Wood’s equation to adjust the experimental data obtained from patient samples collected over several weeks to obtain information from each patient. Such data could allow the association of certain pathologies with the characteristics of the antibody response. The addition of such epidemiological data, combined with the use of artificial intelligence, could provide clues to the possible involvement of the humoral immune response in patient recovery. We selected two models of emerging viral infection (Zika virus and SARS-CoV-2) that have different modes of transmission and clinical manifestations for which Wood’s formula perfectly describes the kinetics of the antibodies induced.

Wood’s equation was first routinely used to follow milk production of cattle [3], a biological process of protein production. It is now commonly used to adjust the kinetics of viraemia [4] and estimate IgG concentrations after vaccination [5]. We used this equation to extrapolate unavailable constants (day when antibody detection becomes positive (pos day), day of maximal response (max day), maximal level of antibody (max level), and day when the antibody detection becomes negative (neg day)) as characteristics of the system studied.

### Patients, materials, and methods

The two SARS-CoV-2-infected patients in this study have been described [6]; the patients, who had mild or moderate disease, were called P1 and P2, respectively. Data for the three ZIKV-infected patients were extracted from a previously published cohort survey [7]. The patients, with or without previous dengue infection, were identified as P3, P4, and P5, respectively. The latter three patients presented with several symptoms of varying duration: a maculopapular rash (11, 3, and 14 days for P3, P4, and P5, respectively), conjunctivitis (12, 3, and 10 days, respectively), pruritis (6 and 3 days for P3 and P4 and not recorded for P5), moderate fever (1 day for both P2 and P4), headache (18 days for P3 only), purpura (P3 only), retroorbital pain (12 days for P3 only), asthenia (17 and 15 days for P3 and P4, respectively), anorexia (P3 only), diarrhea (only P5 for 6 days), arthralgia (knees, ankles, elbows, and wrists for P3 for 18 days and the knees for P4 for 2 days), myalgia (2 days for P3 only), and axial back pain (10, 3, and 2 days for P3, P4, and P5, respectively). The curve fit obtained with the data of each patient was used to calculate the max day and compared to the mean of the max day for the three patients combined.

ELISA using total inactivated ZIKV and recombinant domain III of the ZIKV envelope protein (ZEDIII) was used to determine the IgM and IgG levels, respectively [8]. Anti-SARS-CoV-2 IgG levels were determined using the receptor-binding domain (RBD) of the spike envelope glycoprotein as target [9]. Optical-density ratios (ODr) were calculated by dividing the OD obtained with the target for the same sera with the blank. The antibody levels following infection with ZIKV or a SARS-CoV-2 infection were fitted using the Wood model (ODr=a.Day^b^.exp^(-c.Day)^+d) using KaleidaGraph 4.5 software. The positive threshold of the ODr was calculated as the mean + 3×standard deviation for each studied antibody and antigenic target (IgM for ZIKV=3.00, IgG for EDIII=1.54; IgG for RBD=2.40). Max day was calculated following the formula: max day=b/c. The maximum IgG levels were calculated following the formula: max level =a(b/c)^b^exp^(-b)^ [4]. The Wood curve was plotted day by day for each condition. Both pos day and neg day were extrapolated from each curve.

## Results

The antibody kinetic profile parameters are presented in Table 1. Pos day, max day, max IgG level and neg day were 0, 54, 20.5 and 377 for P1 and 2, 43, 19.3 and 251 days for P2, respectively, for SARS-CoV-2 (Fig 1A). Pos day, max day, max antibody level, and neg day for IgM and IgG were 6, 19, 5.9, and 49, and 3, 153, 5.1 and 680 days, respectively, for P3, who presented no immunological scar, and 6, 14, 3.2, 32 days for IgM and 4, 190, 21.9 and 1,660 days for IgG for P4 (Fig 1B). These data for P5 were 8, 133, 2.4, and 598 days for IgG. The extrapolation of the max day (170 days) and the neg day (832 days) obtained with the curve fit of the pooled data of P3, P4, and P5 (Pooled data) was different from the calculated mean of the max day (159 days) and neg day (979 days) of the three individual curves (Fig 1C). The reliability factors were high (r≥0.88), except for the pooled data curve (r=0.56) (Table 1).

**Table 1.**

| Patient | Age | Sex | Imm. scar | Antibody type | ELISA target | a | b | c | d | r | Pos day | Max day | Max level | Neg day |
| --- | --- | --- | --- | --- | --- | --- | --- | --- | --- | --- | --- | --- | --- | --- |
| P1 | 25-30 | F | ND | IgG | RBD SARS-CoV-2 | 1.513 | 0.87006 | 0.015995 | 1 | 0.93 | 0 | 54 | 20.5 | 377 |
| P2 | 50-55 | M | ND | IgG | RBD SARS-CoV-2 | 0.33971 | 1.4658 | 0.034283 | 1 | 0.88 | 2 | 43 | 19.3 | 251 |
| P3 | 35-40 | M | No | IgM | ZIKV | 0.1585 | 1.8327 | 0.094116 | 1 | 0.97 | 6 | 19 | 5.9 | 49 |
| P3 | 35-40 | M | No | IgG | ZEDIII | 0.017577 | 1.4083 | 0.0092013 | 1 | 0.99 | 3 | 153 | 5.1 | 680 |
| P4 | 40-45 | M | Yes | IgM | ZIKV | 0.39849 | 1.2568 | 0.08709 | 1 | 0.96 | 6 | 14 | 3.2 | 32 |
| P4 | 40-45 | M | Yes | IgG | ZEDIII | 0.10658 | 1.2549 | 0.006617 | 1 | 0.92 | 4 | 190 | 21.9 | 1660 |
| P5 | 40-45 | M | ND | IgG | ZEDIII | 0.12655 | 0.7574 | 0.005674 | 1 | 0.91 | 8 | 133 | 2.4 | 598 |
| Pooled data | None | None | None | IgG | None | 0.0505062 | 1.2854 | 0.0075445 | 1 | 0.56 | 4 | 170 | 10.3 | 832 |

Abbreviations: Imm. scar: immunological scar. a, b, c, d: Wood's constants. r: reliability factor. Pos day: day when antibody detection becomes positive. Max day: day of maximal response. Max level: maximal level of antibody. Neg day: day when the antibody detection becomes negative. ND: Not Documented. F: Female. M: Male. RBD SARS-CoV-2: receptor-binding domain of severe acute respiratory syndrome coronavirus 2. ZIKV: Zika virus. ZEDIII: recombinant domain III of the ZIKV envelope protein.

**Figure 1.**
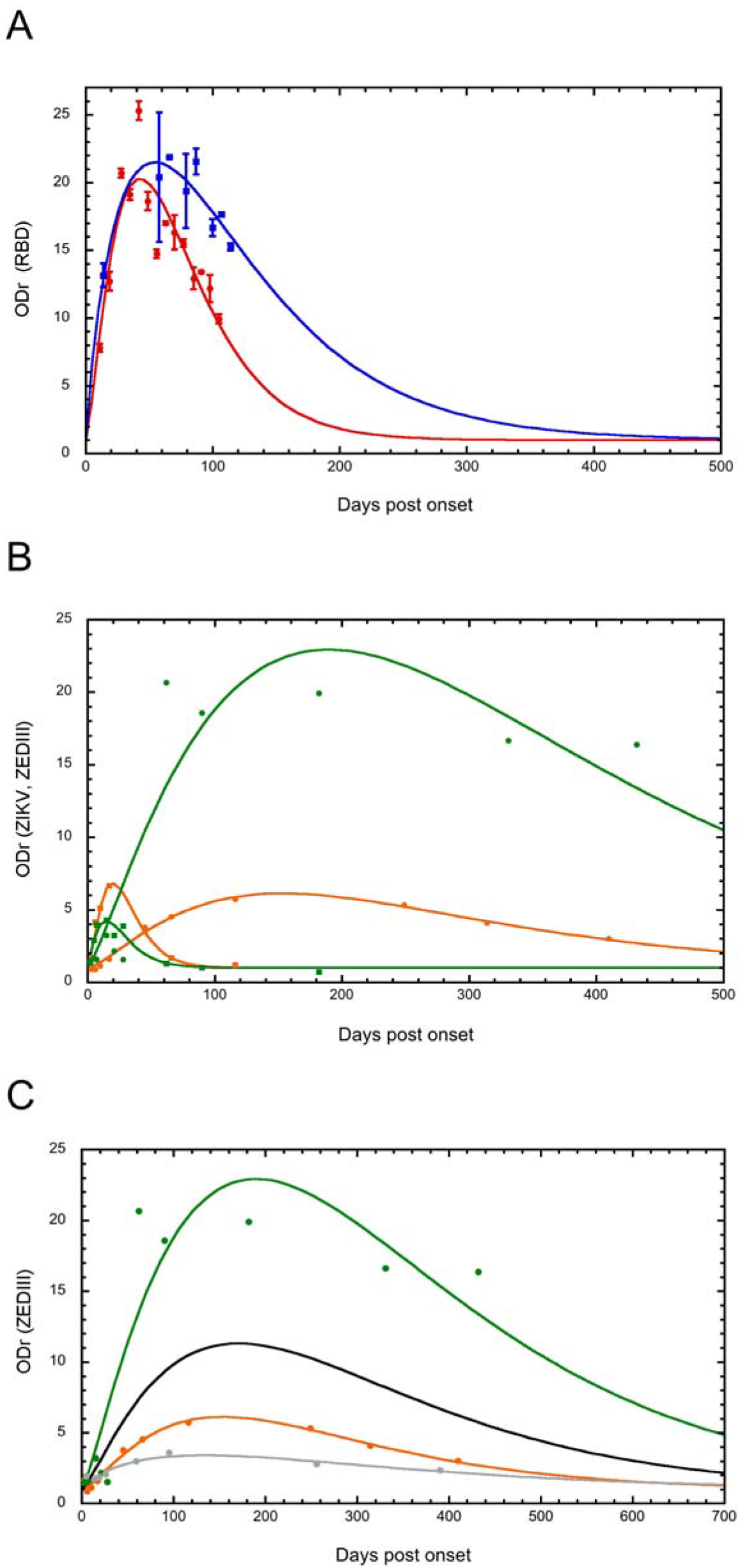
Antibody kinetics. **(A)** IgG of two SARS-CoV-2-infected patients, P1 (blue) and P2 (red) and **(B)** IgM (square) and IgG (circle) of ZIKV-infected patients P3 (orange) and P4 (green) were fitted using the Wood equation. **(C)** The data from three patients were plotted and the curve fit performed using the Wood equation for each patient (P3: orange, P4: green, P5: grey). The black curve is the fitted curve using pooled data of the three patients P1, P2, and P3. The IgM curve reliability factors r are 0.97 and 0.96 for P3 and P4, respectively, and those of the IgG curve 0.93, 0.88, 0.99, 0.92, 0.91, and 0.56 for P1, P2, P3, P4, P5 and the black curve, respectively. The means and standard deviations of the optical density ratios are presented panel A.

## Discussion

ODr values correlate with the concentrations and avidities of antibodies, reflecting their affinity constants and therefore their ability to specifically bind to their target at a determined concentration. Although ODrs are only semi-quantitative, the maximum ODr and determined positivity threshold are intrinsic values of the system and give relevant relative values.

Many samples were missing during the first weeks after the infection of P1 with SARS-CoV-2, but adjustment of the obtained curve gave results close to those of the adjusted curve for P2 (with a high reliability factor, r≥0.88). Samples are rarely taken on pos day or max day and that for neg day is often too late to be taken, sometimes hundreds of days after the onset of symptoms. However, these missing values can be extrapolated with high reliability (r close to 1). In many studies, incomplete data from patients were pooled to obtain a full fit of the kinetic curve and the characteristic constants calculated using the equation obtained by fitting the aggregate data [10, 11].

The means of the max and neg days obtained from the curve fit of each patient (r≥0.91) were very different from the determined max and neg days obtained from a single curve fit (low r=0.56) of the pooled data (Figure 1C). This leads to the loss of information and the ability to observe distinct populations and, finally, to a bias in the estimation of the kinetic parameters, as the immune response varies between patients; here, according to the immune status of the scar vis-à-vis the flavivirus. We propose a method that has already been proven for other biological variables to obtain individual information rather than by pooling the data. Wood’s model makes it possible to adjust the kinetics of each patient and then individually extract each constant.

We applied this method to the humoral immune response directed against two viruses that have different modes of transmission and clinical manifestations. This method allows patients to be linked to a past event, visible, for example, by the presence of a flavivirus immune scar. The patient who was previously infected with a flavivirus had a lower level of IgM directed against ZIKV and a higher level of IgG than the patient without a serological scar directed against a flavivirus, as observed in a previous study [12]. The patient with a serological scar to flavivirus showed much less symptoms than the patient without such a scar. During SARS-CoV-2 infection, the negativation of IgG directed against the RBD domain occurred faster for patient P2, who had more serious symptoms than patient P1, who had a mild form of the disease.

The identification of different kinetic profiles for patients would make it possible to relate a typical profile to the seriousness of the clinical signs and could be useful in predicting the intensity or evolution of the pathology and perhaps even in demonstrating, a posteriori, the association of a type of humoral immune response to improvement or worsening of the patient’s condition. Second, such identification could contribute to the exploration of the mechanisms involved in severe forms and propose solutions for treating patients identified to have a similar kinetic profile. The diagnostic window could also be determined and would be useful for the diagnosis of diseases such as dengue-like syndrome or Covid-19.

In conclusion, we present a methodology that makes it possible to obtain otherwise unavailable individual data. These data could help identify patients with identical profiles and thus be useful in classifying immune responses associated with disease severity, highlighting mechanisms that are hidden in pooled data.

## Data Availability

all data are available in the manuscript

## Acknowledgements

This work was partially supported by the European Union’s Horizon H2020 Project “Advanced Nanosensing platforms for Point of care global diagnostics and surveillance” (CONVAT) (No101003544) and partially funded by the Direction Générale de l’Armement and the Service de Santé des Armées (Biomedef PDH-2-NRBC-2-B-2111), and the Direction centrale du Service de santé des armées (grant IDs: 2016RC10).

